# Domain-based basal and daytime ambulatory glycemic exposure metrics derived from continuous glucose monitoring: a real-world clinic-based study

**DOI:** 10.64898/2026.05.24.26353983

**Authors:** Suresh N. Shinde, Rituparna S. Shinde, Succhietra Y. Bhangaaley

## Abstract

**Background:** Consensus continuous glucose monitoring (CGM) metrics, including time in range (TIR), time above range (TAR), time below range (TBR), mean glucose, glucose management indicator, and glycemic variability, are essential for modern glucose assessment. However, these whole-day summaries do not explicitly partition nocturnal basal from daytime ambulatory glycemic burden.

**Objective:** To develop and evaluate a complementary domain-based CGM framework that quantifies basal and daytime ambulatory glycemic exposure across oral glucose tolerance test (OGTT)-derived dysglycemia phenotypes.

**Methods:** In this observational, clinic-based study, 253 individuals underwent OGTT with insulin measurement and CGM. Participants were classified using a prespecified OGTT-derived phenotyping algorithm, implemented through a deterministic rules-based web calculator, and collapsed into five groups: NoDM, Increased insulin resistance, Midzone Glycemia, Prediabetes, and Diabetes. CGM files were uniformly reprocessed by selecting the latest contiguous episode and retaining the most recent 15 calendar days with data. The 24-hour profile was partitioned into nocturnal basal (00:00 to <06:00) and daytime ambulatory (06:00 to <24:00) domains. Derived indices included Area of Basal Glycemia (ABG), Area of Prandial/Daytime Ambulatory Glycemia (APG), incremental ABG (iABG), incremental APG (iAPG), and exploratory deficit indices dABG and dAPG.

**Results:** The final dataset contributed 3,647 analyzable CGM days. APG remained higher than ABG across all groups. Mean ABG/APG increased from 80.45/86.38 mg/dL in NoDM to 111.96/124.70 mg/dL in Diabetes. Mean iABG/iAPG increased from 5.65/6.60 to 34.12/38.91 mg/dL, whereas dABG/dAPG declined as dysglycemia worsened.

**Conclusions:** The ABG/APG framework provides interpretable, domain-resolved CGM burden metrics that separate basal from daytime ambulatory exposure and distinguish total burden from above-threshold excess. These indices are proposed as adjunctive metrics to support dysglycemia phenotyping, early risk recognition, and treatment monitoring, but are not intended to replace established consensus CGM metrics or diagnostic criteria. External, prospective validation is required.

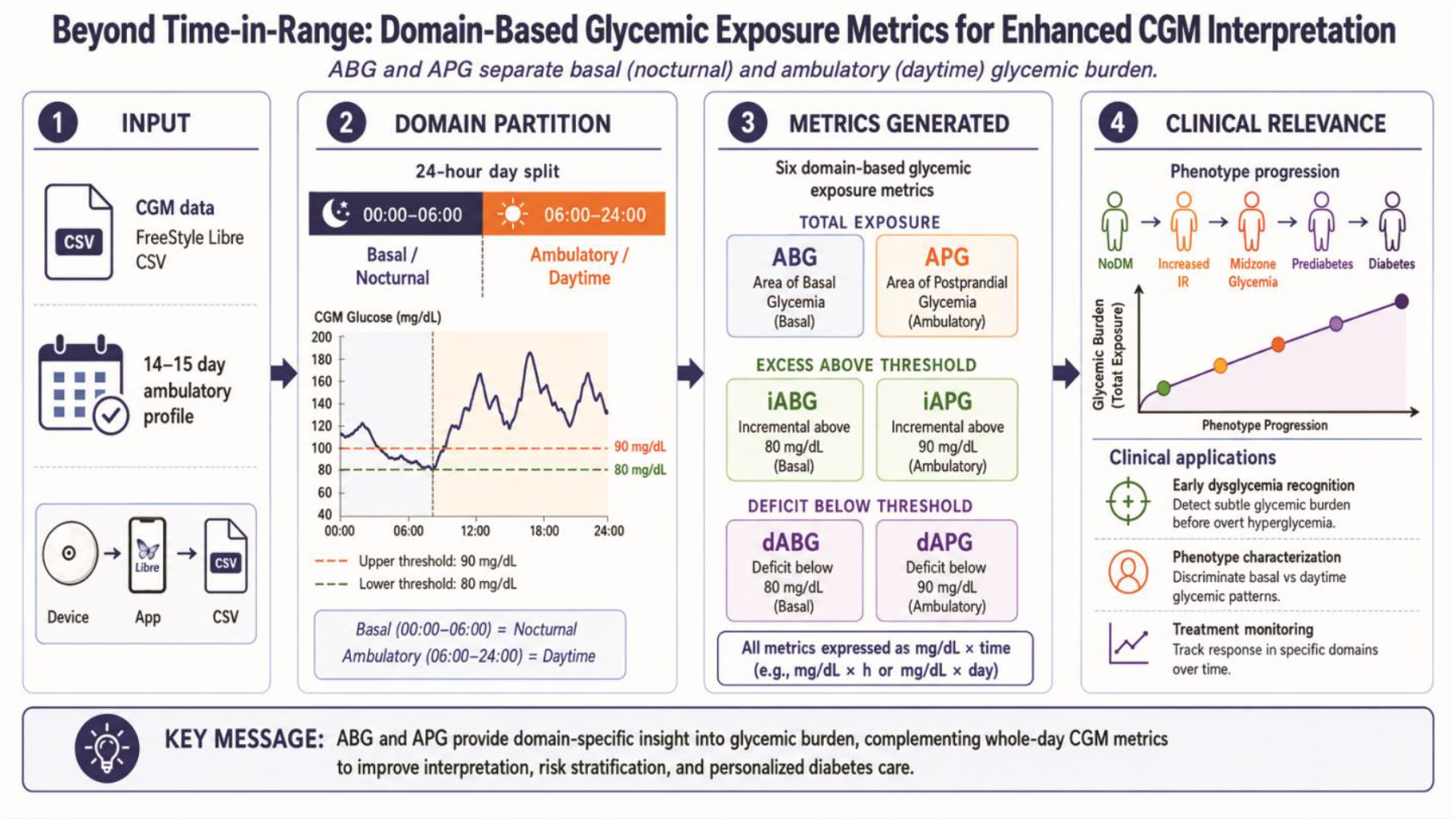

## Introduction

Continuous glucose monitoring (CGM) has transformed glycemic assessment by shifting from isolated fasting, post-load, and glycated hemoglobin measurements to continuous characterization of glucose exposure, including time in range (TIR), time above range (TAR), time below range (TBR), glucose management indicator (GMI), and glycemic variability [1-8]. These measures have become central to modern diabetes care and are increasingly used in research settings involving prediabetes, obesity, cardiometabolic risk, pregnancy, dietary interventions, and monitoring of pharmacologic therapy. The framework proposed here is intended to complement, not replace, these established CGM metrics.

Despite this progress, most widely used CGM metrics summarize glucose behavior over the entire 24-hour period. They therefore do not explicitly indicate whether glycemic burden is concentrated in the nocturnal basal domain or the daytime ambulatory domain. This distinction may be important because meals less influence overnight glucose and may more closely reflect basal hepatic glucose output, insulin sensitivity, counter-regulatory tone, and sleep-related physiology. Daytime glucose exposure, in contrast, reflects a composite free-living state shaped by basal glycemia, nutrient absorption, meal timing, physical activity, circadian effects, peripheral glucose disposal, medications, and behavioral variability.

Two individuals may therefore have similar whole-day mean glucose, GMI, TIR, or glycemia risk summaries while differing substantially in the temporal location and physiological meaning of the burden. A domain-resolved exposure framework may add interpretive value by identifying whether the predominant burden is nocturnal, daytime, above-threshold, or below-threshold.

To address this interpretive gap, we developed a set of CGM-derived, domain-based exposure indices. Area of Basal Glycemia (ABG) quantifies time-normalized glucose exposure during the nocturnal basal window from 00:00 to <06:00 hours. Area of Prandial Glycemia (APG), retained as the acronym used in our prior implementation, quantifies daytime ambulatory exposure from 06:00 to <24:00 hours and should be interpreted as a composite daytime ambulatory burden metric rather than a pure meal-excursion metric. Incremental ABG (iABG) and incremental APG (iAPG) isolate the excess burden above threshold, while deficit indices dABG and dAPG quantify the mirrored below-threshold burden.

The present study evaluates these indices in a clinic-based cohort in which all participants underwent both OGTT with insulin measurement and CGM. The primary aim was to determine whether ABG, APG, iABG, and iAPG vary consistently across OGTT-derived dysglycemia phenotypes. Secondary aims were to examine the behavior of dABG and dAPG, relate the CGM-derived burden indices to OGTT glucose area under the curve (AUC-glucose), and describe a browser-based implementation pathway for clinic-level deployment.

## Methods

### Study design and ethics

This observational study was conducted within a metabolic phenotyping program in which participants underwent OGTT-based metabolic evaluation and CGM. The analytical objective was to assess whether domain-based CGM-derived burden indices track the progression of dysglycemia when interpreted against an OGTT-derived phenotypic scaffold.

The Ethics Committee of Poona Hospital and Research Center, Pune, India, reviewed the study protocol. The Committee granted an ethics-review waiver for the observational study titled ‘To develop and evaluate a domain-based CGM framework that quantifies basal and ambulatory glycemic exposure across OGTT-derived dysglycemia groups’ (Ref. No. RECH/EC/2026-27/049; dated 19 May 2026). The analysis used clinic-derived observational data and did not involve any study-specific intervention.

### Participants and source data

The final analytic cohort comprised 253 individuals with paired OGTT and CGM data. Each participant underwent OGTT with glucose and insulin measurements as part of metabolic phenotyping. CGM data were obtained using FreeStyle Libre systems and uniformly reprocessed before calculation of the proposed indices.

Broad dysglycemia categories were assigned using an OGTT-derived phenotypic framework and defined as NoDM, Increased insulin resistance (Increased IR), Midzone Glycemia, Prediabetes, and Diabetes. These categories were used to preserve a clinically meaningful continuum of dysglycemia while avoiding excessive expansion of phenotype definitions in the main manuscript.

### OGTT-derived phenotype classification and web-app implementation

OGTT-derived phenotypes were assigned using a pre-specified, rule-based classification algorithm implemented in the OGTT-Plus web calculator, developed by the study group and hosted at ‘www.OGTT.in’, during the analysis workflow. The web application served as a deterministic implementation layer, applying fixed glucose- and insulin-based phenotype rules uniformly across records. It was not used as an adaptive, machine-learning, or autonomous diagnostic decision-making system.

In the present analysis, the detailed OGTT output categories were collapsed into five broad groups. NoDM denoted participants without OGTT-defined diabetes or prediabetes and without an intermediate glycemic excursion above the midzone threshold. Increased IR denoted participants with nondiabetic glucose values but an elevated insulin-resistance signal within the algorithm. Midzone Glycemia was defined as fasting glucose <100 mg/dL and 120-minute glucose <140 mg/dL, with intermediate OGTT glucose >155 mg/dL at the 30-minute, 60-minute, or both 30- and 60-minute time points. Prediabetes included impaired fasting glucose, impaired glucose tolerance, or their combination. Diabetes included participants who met diabetes-range OGTT criteria or had a known diabetes status, as captured in the clinical dataset.

The OGTT-Plus implementation ensured consistent application of the same classification logic to all records. The phenotype classification rules and the analysis-locked OGTT-Plus calculator version used for this study were archived and are supplied in Supplementary Methods 1 (Supplemental 1). The web calculator served only as a deterministic implementation layer for applying fixed, pre-specified phenotype rules uniformly across the dataset.

### CGM preprocessing

All available raw CGM files for included participants were sorted chronologically. CGM episodes were defined whenever the interval between consecutive readings exceeded 24 hours. The latest contiguous episode was selected, and within that episode, only the most recent 15 calendar days with data were retained.

To ensure representation of both physiological domains, the analytic inclusion criteria required at least 5 valid nocturnal basal days and at least 5 valid daytime ambulatory days. After reprocessing and eligibility filtering, the final strict analytic CGM dataset comprised 253 participants contributing 3,647 analyzable CGM days.

Because sampling density may vary across FreeStyle Libre versions, CGM traces were harmonized to a common analytical structure before burden calculation. The analytical objective was comparability of time-integrated burden indices across participants rather than preserving native device-specific sampling density.

### Definition of basal and daytime ambulatory domains

The 24-hour day was divided into two fixed domains: a nocturnal basal window from 00:00 to <06:00 hours and a daytime ambulatory window from 06:00 to <24:00 hours. The nocturnal window was selected to represent the relatively stable basal portion of the day, less directly influenced by meal-related excursions. The daytime window was intended to capture composite ambulatory glycemic exposure arising from basal glycemia, nutrient absorption, meal-related excursions, activity, circadian influences, peripheral glucose disposal, and treatment effects.

The physiological reference anchors for incremental and deficit indices were derived from a predefined, strict NoDM reference subset within the study framework. The nocturnal basal domain was referenced to 80 mg/dL, and the daytime ambulatory domain to 90 mg/dL.

### Derivation of CGM burden indices

Let G(t) denote the interstitial glucose concentration at time t. For each analyzable day, glucose exposure within each domain was quantified using trapezoidal integration and normalized by the duration of the corresponding window.Therefore, all indices are time-normalized AUC-based measures reported in mg/dL.

For the nocturnal basal window, ABG = (1/6) x integral G(t) dt from 00:00 to <06:00 hours. For the daytime ambulatory window, APG = (1/18) x integral G(t) dt from 06:00 to <24:00 hours.

To isolate excess burden above the threshold, incremental indices were derived using threshold-truncated integration. For the nocturnal domain, iABG = (1/6) x integral max[G(t) - 80, 0] dt from 00:00 to <06:00 hours. For the daytime domain, iAPG = (1/18) x integral max[G(t) - 90, 0] dt from 06:00 to <24:00 hours. Negative values were truncated to zero before integration.

As exploratory mirrored extensions, deficit indices quantified below-threshold burden. For the nocturnal domain, dABG = (1/6) x integral max[80 - G(t), 0] dt from 00:00 to <06:00 hours. For the daytime domain, dAPG = (1/18) x integral max[90 - G(t), 0] dt from 06:00 to <24:00 hours. These deficit metrics were treated as exploratory rather than primary endpoints.

### ABG/APG calculator and software implementation

The study group developed a browser-based ABG/APG Companion application as the practical implementation layer for the CGM analytics workflow. The tool imports standard FreeStyle Libre CSV export files, identifies the latest contiguous analyzable episode, retains the most recent valid CGM days for analysis, partitions each day into nocturnal basal and daytime ambulatory domains, calculates ABG, APG, iABG, iAPG, dABG, dAPG, TAR, and TBR summaries using pre-specified formulae, and generates a printable clinical report.

The ABG/APG Companion application served as a deterministic calculation and reporting interface. The present study evaluates the behavior of the proposed indices and their relationship to OGTT-derived phenotypes; it does not constitute validation of the application as a regulated medical device, autonomous diagnostic software, or software-as-a-medical-device product.

### Statistical analysis

The primary variables of interest were ABG, APG, iABG, and iAPG. Secondary variables included dABG, dAPG, HbA1c, mean CGM glucose, and OGTT-derived AUC glucose. Continuous variables were summarized as mean ± standard deviation or as median with interquartile range, and categorical variables as counts and percentages.

Phenotype-wise comparisons were interpreted descriptively across the ordered dysglycemia spectrum. Trends were evaluated both graphically and by inspection of grouped means. Data curation, index computation, and figure generation were performed using spreadsheet-based workflows, Python-based analytical tools, and the browser-based implementation described above.

## Results

### Cohort characteristics and phenotype distribution

The final analytic CGM dataset comprised 253 participants contributing 3,647 analyzable CGM days. Cohort characteristics and the distribution of grouped phenotypes are summarized in Table 1. The mean age was 46.2 ± 10.4 years, and the cohort included 153 men and 100 women. Participants contributed an average of 14.4 ± 1.3 CGM days, with a median of 15 days (IQR 14-15). The mean HbA1c was 6.79 ± 1.66%, and the mean CGM glucose was 109.5 ± 32.7 mg/dL.

**Table 1.** Cohort characteristics and grouped phenotype distribution.

| Section | Characteristic | Value |
| --- | --- | --- |
| Overall cohort | Participants, n | 253 |
| Overall cohort | Analyzable CGM-days, n | 3,647 |
| Overall cohort | Age, mean ± SD (years) | 46.2 ± 10.4 |
| Overall cohort | Sex, male/female | 153/100 |
| Overall cohort | CGM days per participant, mean ± SD | 14.4 ± 1.3 |
| Overall cohort | CGM days per participant, median (IQR) | 15 (14-15) |
| Overall cohort | HbA1c, mean ± SD (%) | 6.79 ± 1.66 |
| Overall cohort | Mean CGM glucose, mean ± SD (mg/dL) | 109.5 ± 32.7 |
| Grouped phenotype distribution | NoDM | 9 (3.6%) |
| Grouped phenotype distribution | Increased IR | 7 (2.8%) |
| Grouped phenotype distribution | Midzone Glycemia | 25 (9.9%) |
| Grouped phenotype distribution | Prediabetes | 78 (30.8%) |
| Grouped phenotype distribution | Diabetes | 134 (53.0%) |

Using the OGTT-derived grouped phenotype framework, 9 participants (3.6%) were classified as NoDM, 7 (2.8%) as Increased IR, 25 (9.9%) as Midzone Glycemia, 78 (30.8%) as Prediabetes, and 134 (53.0%) as Diabetes. Thus, the cohort spanned the dysglycemia spectrum but was weighted toward later stages.

### Physiological reference subset

The strict NoDM subset served as the physiological reference group (Table 2). In this subset, mean ABG was 80.45 ± 9.33 mg/dL, mean APG was 86.38 ± 11.39 mg/dL, mean iABG was 5.65 ± 5.28 mg/dL, and mean iAPG was 6.60 ± 5.68 mg/dL. Mean dABG and dAPG were 3.91 ± 3.57 mg/dL and 5.63 ± 4.82 mg/dL, respectively. These values indicate that, in metabolically normal participants, both basal and daytime ambulatory glycemic domains remained close to physiological reference levels, while the excess above-threshold burden remained low.

**Table 2.** Grouped phenotype means for CGM burden indices and OGTT AUC glucose.

| Grouped phenotype | n | ABG mean ± SD | APG mean ± SD | iABG mean ± SD | iAPG mean ± SD | dABG mean ± SD | dAPG mean ± SD | AUC glucose mean |
| --- | --- | --- | --- | --- | --- | --- | --- | --- |
| NoDM | 9 | 80.45 ± 9.33 | 86.38 ± 11.39 | 5.65 ± 5.28 | 6.60 ± 5.68 | 3.91 ± 3.57 | 5.63 ± 4.82 | 222.7 |
| Increased IR | 7 | 81.41 ± 8.71 | 87.81 ± 12.75 | 5.53 ± 5.20 | 6.02 ± 7.13 | 3.34 ± 3.95 | 5.71 ± 4.58 | 220.8 |
| Midzone Glycemia | 25 | 81.20 ± 10.63 | 88.21 ± 10.63 | 6.43 ± 7.03 | 7.39 ± 6.18 | 4.12 ± 4.32 | 6.53 ± 4.75 | 297.1 |
| Prediabetes | 78 | 87.25 ± 12.55 | 92.40 ± 12.94 | 10.93 ± 10.15 | 10.16 ± 9.02 | 2.86 ± 3.45 | 5.08 ± 4.42 | 336.3 |
| Diabetes | 134 | 111.96 ± 35.52 | 124.70 ± 36.93 | 34.12 ± 34.55 | 38.91 ± 35.33 | 1.42 ± 2.67 | 1.90 ± 3.15 | 530.5 |

### Total basal and daytime ambulatory glycemic burden across phenotypes

Grouped phenotype means for the CGM burden indices are shown in Table 2 and illustrated in Figure 1. Mean ABG values across NoDM, Increased IR, Midzone Glycemia, Prediabetes, and Diabetes were 80.45, 81.41, 81.20, 87.25, and 111.96 mg/dL, respectively. Corresponding mean APG values were 86.38, 87.81, 88.21, 92.40, and 124.70 mg/dL, respectively.

**Figure 1.**
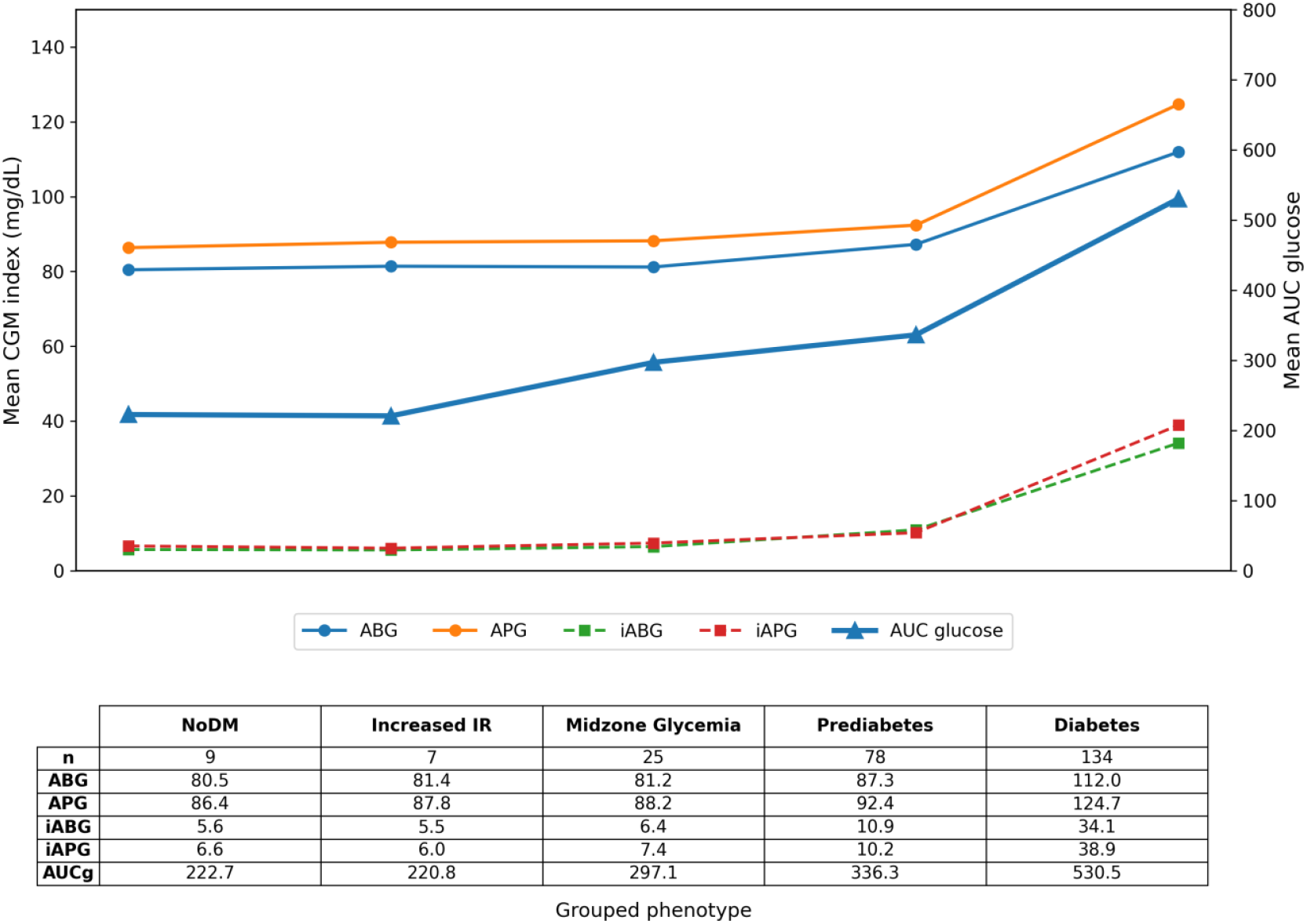
Domain-based CGM burden indices across OGTT-derived dysglycemia phenotypes. The figure summarizes grouped trends in ABG, APG, iABG, iAPG, dABG, dAPG, and OGTT AUC glucose across NoDM, Increased insulin resistance, Midzone Glycemia, Prediabetes, and Diabetes. ABG, Area of Basal Glycemia; APG, Area of Prandial/Daytime Ambulatory Glycemia; iABG/iAPG, incremental above-threshold basal/daytime burden; dABG/dAPG, below-threshold deficit burden; AUC, area under the curve; OGTT, oral glucose tolerance test.

APG remained consistently higher than ABG across all grouped phenotypes. Both total-burden indices were relatively compact in earlier dysglycemia groups and rose more sharply in diabetes, suggesting that overt diabetes is associated with an expansion of glycemic burden across both daytime and nocturnal domains.

### Incremental and deficit burden indices

The incremental indices showed the clearest widening across grouped phenotypes (Table 2; Figure 1). Mean iABG values were 5.65, 5.53, 6.43, 10.93, and 34.12 mg/dL for NoDM, Increased IR, Midzone Glycemia, Prediabetes, and Diabetes, respectively. Mean iAPG values were 6.60, 6.02, 7.39, 10.16, and 38.91 mg/dL, respectively. Compared with the total-burden indices, the excess-burden indices showed sharper separation in later dysglycemia, particularly in diabetes.

In contrast, deficit-burden signals were generally higher in non-diabetic and early dysglycemic states and progressively attenuated as dysglycemia worsened. Mean dABG values were 3.91, 3.34, 4.12, 2.86, and 1.42 mg/dL across the five groups, whereas mean dAPG values were 5.63, 5.71, 6.53, 5.08, and 1.90 mg/dL, respectively. These findings suggest that below-threshold burden is progressively replaced by sustained above-threshold exposure as dysglycemia advances.

### Relationship with OGTT-derived glycemic burden

The grouped phenotype trend plot in Figure 1 shows that the CGM-derived burden indices remain coherent when interpreted against OGTT AUC glucose as the glycemic reference. Across the five broad dysglycemia stages, AUC glucose rose progressively, with the greatest increase observed in diabetes. The CGM indices broadly tracked this progression. APG showed the strongest overall-burden rise across the grouped spectrum, while iABG and iAPG showed the clearest expansion of above-threshold burden.

Taken together, these findings show that ABG, APG, iABG, and iAPG vary systematically across OGTT-derived grouped dysglycemia phenotypes. APG remained consistently higher than ABG, confirming a greater daytime ambulatory burden than basal-domain burden across the spectrum. The clearest pathological widening was observed in iABG and iAPG, especially in diabetes, whereas dABG and dAPG decreased as dysglycemia worsened.

### Digital implementation

The ABG/APG Companion application provided a deterministic implementation of the analytical workflow, including CGM import, episode selection, time-domain segmentation, burden-index calculation, TAR/TBR summary generation, and printable report output. Figure 2 shows a representative anonymized report generated by the application from FreeStyle Libre historical glucose data. This software layer enables reproducible clinic-level deployment of the framework while preserving the formula-based transparency of the indices.

**Figure 2.**
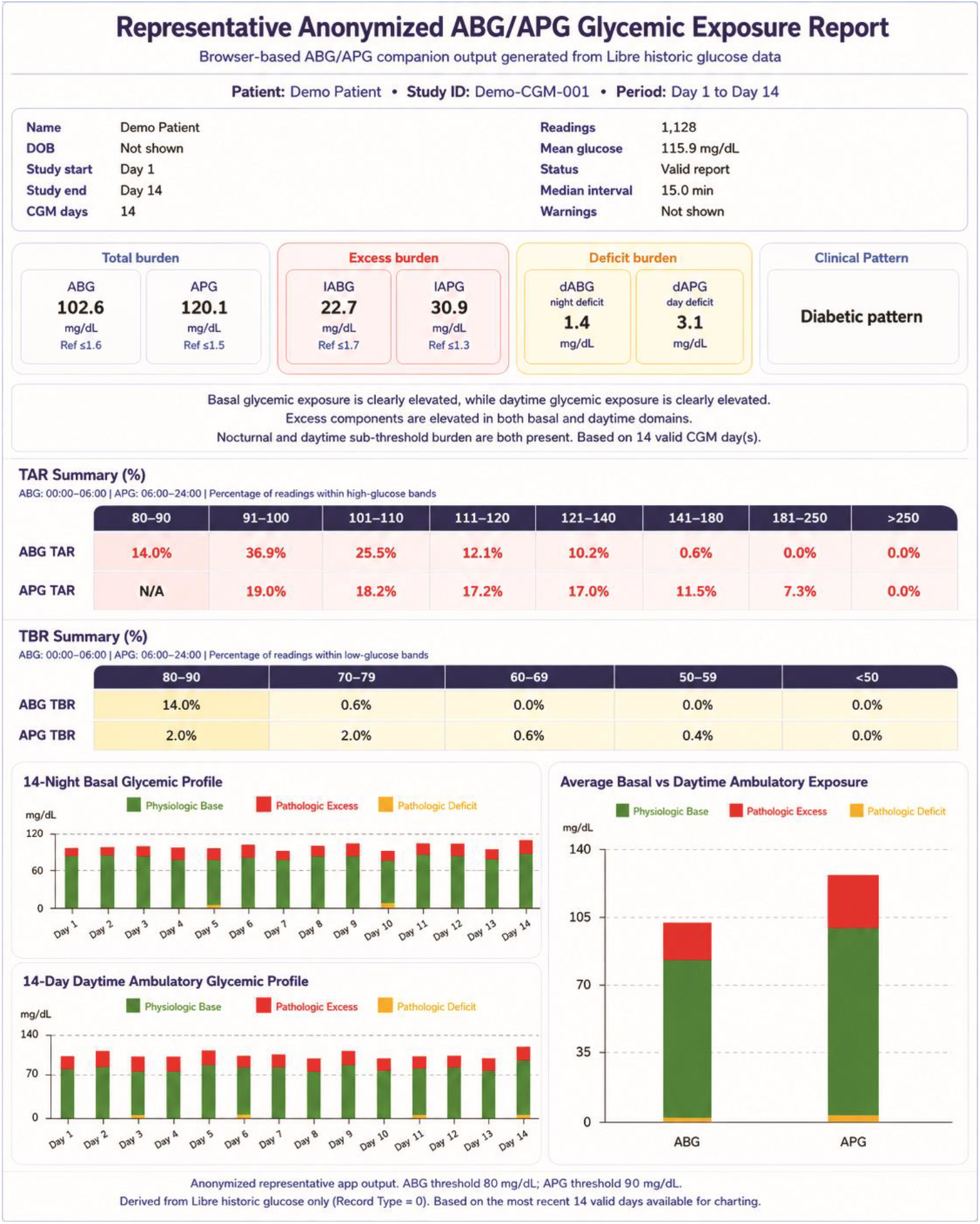
Representative ABG/APG Companion report generated from FreeStyle Libre historical glucose data. The browser-based application imports standard CGM exports, selects the latest contiguous analyzable episode, segments the 24-hour profile into nocturnal basal and daytime ambulatory domains, calculates domain-wise total, incremental, and deficit burden indices, summarizes TAR/TBR bins, and generates a printable report. The figure is illustrative and does not imply validation of the application as autonomous diagnostic software.

## Discussion

In this study, we evaluated a domain-based CGM framework that partitions 24-hour glucose exposure into a nocturnal basal domain and a daytime ambulatory domain, and further divides each domain into total burden, above-threshold excess burden, and below-threshold deficit burden. The principal finding is that ABG, APG, iABG, and iAPG vary systematically across five OGTT-derived categories of dysglycemia, with the clearest separation observed in the incremental above-threshold components and the greatest absolute burden in diabetes.

A major conceptual strength of the ABG/APG framework is that it provides a complementary domain-resolved layer alongside currently used whole-day CGM outputs. Contemporary CGM reporting emphasizes mean glucose, GMI, TIR, TAR, TBR, glycemic variability metrics, and risk-oriented summaries [1-9]. These measures are clinically valuable, but they do not explicitly distinguish between the nocturnal basal and daytime ambulatory domains. ABG and APG localize glycemic exposure to physiologically distinct time domains, while iABG and iAPG quantify excess above physiologic anchors and dABG/dAPG capture mirrored low-side burden.

This distinction is clinically relevant because similar whole-day CGM summaries do not necessarily indicate similar exposure burden. Two individuals may have similar TIR yet differ in the magnitude, timing, and domain distribution of glycemic burden. The same logic applies to GMI, mean glucose, GRI, and variability summaries: each provides useful whole-day information, but none localizes burden as ABG/APG do. Thus, the proposed framework should be viewed as a domain-resolved, exposure-based extension of current CGM metrics rather than a replacement for them.

The grouped phenotype data support the physiological rationale for this partitioning. ABG is intended to reflect integrated nocturnal basal glycemia, a relatively stable domain less directly influenced by meals. APG captures the broader daytime ambulatory burden arising from basal glycemia, nutrient absorption, excursions, activity, circadian influences, peripheral disposal, and treatment effects. Across the spectrum of grouped dysglycemia, the consistent excess of APG over ABG is biologically plausible and supports interpreting APG as the stronger metric of integrated daytime burden.

The relationship between the grouped burden indices and OGTT AUC glucose strengthens the framework’s translational interpretation. Because all included participants underwent OGTT with insulin estimation, the CGM-derived indices could be interpreted against a metabolically characterized scaffold rather than against CGM alone. APG broadly paralleled worsening AUC glucose across groups, whereas ABG rose more modestly initially and then more clearly in later dysglycemia. One coherent interpretation is that, as dysglycemia worsens, daytime ambulatory burden progressively extends into the nocturnal basal domain, producing a form of nocturnal spillover. This should be regarded as a conceptual interpretation rather than a validated threshold phenomenon.

The incremental indices add an interpretive layer by separating physiological glucose exposure from the excess burden above the threshold. Total ABG and APG include physiological glycemia, whereas iABG and iAPG quantify only the burden above the reference anchors. This distinction may be useful when comparing early dysglycemia, prediabetes, and diabetes, because the incremental indices more clearly highlight pathological widening than total-burden indices alone.

A practical strength of the framework is its digital deployability. The ABG/APG Companion application implements the analytical pipeline as a browser-based tool that imports standard CGM exports, performs episode selection and time-domain segmentation, calculates domain-wise burden indices, generates TAR/TBR summaries, and produces a printable report. This implementation demonstrates that the metrics are computationally lightweight and can be deployed from standard CGM exports. However, the present study evaluates the behavior of the metrics rather than validating the application as autonomous diagnostic software or as a regulated medical device.

Recent outcome-validation work has strengthened the clinical relevance of CGM-derived metrics. A contemporary virtual-CGM reanalysis of the Diabetes Control and Complications Trial reported that 14-day CGM metrics, including TIR, mean glucose, time above range, and HBGI, were associated with microvascular outcomes, broadly paralleling HbA1c [10]. Although this evidence comes from type 1 diabetes and does not specifically validate ABG/APG, it supports the broader premise that CGM-derived exposure metrics can provide clinically meaningful information.

The framework may have clinical value for early recognition of dysglycemia and risk stratification. We do not propose CGM alone as a diagnostic test for diabetes or prediabetes because current diagnostic standards remain based on glucose and HbA1c criteria [11]. Instead, ABG/APG may serve as adjunctive CGM-derived burden markers in clinical situations where dysglycemia risk is high but conventional measures may be borderline or discordant, including dysmetabolic obesity, first-degree relatives of people with diabetes, metabolic dysfunction-associated steatotic liver disease, polycystic ovary syndrome, obstructive sleep apnea, gestational diabetes risk, and cardiometabolic risk assessment.

The framework may also be useful for longitudinal treatment monitoring. Because the indices can be calculated on a daily basis, changes in ABG, APG, iABG, and iAPG could potentially quantify early response to dietary intervention, medication adjustment, weight-loss therapy, or structured lifestyle programs. For example, a reduction in early-morning ABG or iABG over a 14-to 15-day CGM episode may serve as a practical marker of improved basal glycemic regulation, whereas a reduction in APG or iAPG may reflect a reduced daytime ambulatory burden. These applications are hypothesis-generating and require prospective validation.

The exploratory deficit indices add a mirrored low-side dimension. dABG and dAPG were higher in non-diabetic and early dysglycemic states and declined with worsening dysglycemia, showing the inverse pattern of the above-threshold indices. This supports the framework’s internal logic: as above-threshold burden expands, below-threshold burden contracts. Their clinical meaning regarding low-glucose exposure, medication titration, and hypoglycemia risk will require independent study.

This study has limitations. It is observational and clinic-based, so the observed burden patterns may not generalize directly to other populations. The cohort was weighted toward diabetes, and the strict NoDM reference subset was small. The physiological anchors were derived within the project framework and require external validation. Using broad OGTT-derived groups preserved clarity but compressed within-group heterogeneity. The web calculators used in the workflow were deterministic implementation tools, but the calculator interfaces themselves have not been independently validated as medical devices. Finally, the indices have not yet been prospectively benchmarked against incident diabetes, microvascular outcomes, cardiovascular events, or intervention response.

In conclusion, the ABG/APG framework offers a physiologically interpretable and digitally deployable extension of CGM interpretation by decomposing 24-hour glucose exposure into basal and daytime ambulatory domains and by separating total exposure from above-threshold excess. APG emerged as the stronger integrated daytime ambulatory burden metric, while ABG provided complementary nocturnal basal information. The incremental indices most clearly highlighted dysglycemia progression. These findings support further external and prospective validation of domain-resolved CGM burden metrics as adjunctive tools for dysglycemia phenotyping and metabolic monitoring.

## Data Availability

All data produced in the present study are available upon reasonable request to the authors.

## Abbreviations

ABG: Area of Basal Glycemia
APG: Area of Prandial/Daytime Ambulatory Glycemia
AUC: area under the curve
CGM: continuous glucose monitoring
GMI: glucose management indicator
iABG: incremental ABG
iAPG: incremental APG
dABG: deficit ABG
dAPG: deficit APG
OGTT: oral glucose tolerance test
TIR: time in range.

## Declarations

### Ethics approval

The Ethics Committee of Poona Hospital and Research Centre, Pune, India reviewed the study protocol. The Committee granted an ethics review waiver for this observational study (Ref. No. RECH/EC/2026-27/049; dated 19 May 2026).

### Data availability

The datasets analyzed in the present study contain clinic-derived health information and are not publicly available. De-identified aggregate data and additional methodological details may be provided by the corresponding author upon reasonable request, subject to institutional and ethical constraints.

### Software and algorithm availability

The OGTT-Plus phenotype calculator hosted at www.OGTT.in and the ABG/APG Companion application were developed by the study group as browser-based, deterministic implementation tools. The phenotype rules and the analysis-locked OGTT-Plus calculator version used in this analysis are archived and provided as Supplementary Methods 1. The ABG/APG formulae are fully specified in the Methods section. De-identified aggregate data and additional implementation details are available from the corresponding author upon reasonable request, subject to institutional and ethical restrictions.

### Author contributions

S.N.S. conceptualized the ABG/APG framework, developed the clinical and analytical hypotheses, supervised data curation, and drafted the manuscript. R.S.S. contributed to clinical interpretation, manuscript review, and cardiometabolic framing. S.Y.B. contributed to clinical data coordination, curation support, and manuscript review. All authors reviewed and approved the final manuscript.

## Acknowledgements

The authors acknowledge the clinical and technical support teams involved in CGM data acquisition, data curation, and calculator implementation.

## Supplementary Methods 1. OGTT Phenotype Classification Rules and Calculator Version Archive

### S1.1 Overview

OGTT-derived phenotypes were assigned using a pre-specified, deterministic rules-based algorithm implemented in the OGTT-Plus web calculator developed by the study group. The algorithm used plasma glucose values at 0, 30, 60, and 120 minutes, together with insulin-derived indices where applicable. The calculator applied fixed logical rules and did not use adaptive modeling, machine learning, or automated diagnostic prediction.

For the present analysis, detailed OGTT-derived categories were collapsed into five broad ordered categories: NoDM, Increased Insulin Resistance, Midzone Glycemia, Prediabetes, and Diabetes.

### S1.2 Broad phenotype rules

| Broad category | Rule used in the present analysis |
| --- | --- |
| NoDM | Fasting plasma glucose <100 mg/dL, 120-minute glucose <140 mg/dL, 30-minute glucose <=155 mg/dL, 60-minute glucose <=155 mg/dL, and no insulin-resistance flag according to the pre-specified insulin-derived criterion. |
| Increased Insulin Resistance | Fasting plasma glucose <100 mg/dL, 120-minute glucose <140 mg/dL, 30-minute glucose <=155 mg/dL, 60-minute glucose <=155 mg/dL, with increased insulin resistance according to the pre-specified insulin-derived criterion used in the OGTT-Plus calculator. |
| Midzone Glycemia | Fasting plasma glucose <100 mg/dL and 120-minute glucose <140 mg/dL, with intermediate glucose >155 mg/dL at the 30-minute time point, the 60-minute time point, or at both 30- and 60-minute time points. |
| Prediabetes | Impaired fasting glucose, impaired glucose tolerance, or both, without meeting criteria for diabetes. |
| Diabetes | Fasting plasma glucose >=126 mg/dL and/or 120-minute glucose >=200 mg/dL, or known diabetes status as captured in the clinical dataset. |

### S1.3 Detailed midzone-glycemia subcategories

| Detailed phenotype | Rule |
| --- | --- |
| 30-minute Midzone Hyperglycemia | Fasting glucose <100 mg/dL, 120-minute glucose <140 mg/dL, 30-minute glucose >155 mg/dL, and 60-minute glucose <=155 mg/dL. |
| 60-minute Midzone Hyperglycemia | Fasting glucose <100 mg/dL, 120-minute glucose <140 mg/dL, 30-minute glucose <=155 mg/dL, and 60-minute glucose >155 mg/dL. |
| 30- and 60-minute Midzone Hyperglycemia | Fasting glucose <100 mg/dL, 120-minute glucose <140 mg/dL, and both 30-minute and 60-minute glucose values >155 mg/dL. |

### S1.4 Calculator version archive

The phenotype classification was performed using an analysis-locked version of the OGTT-Plus calculator hosted at ogtt.in during the study period. For reproducibility, the version used for this analysis was archived with the following submission label:

Calculator name: OGTT-Plus Phenotype Calculator

Analysis-locked version label: OGTT-Plus v2026.05.23-analysis

Hosting location during analysis: ogtt.in

Function: Deterministic OGTT phenotype classification using fixed glucose and insulin-derived rules Analysis dataset: CGM-OGTT analytic cohort, n=253

Output categories used in manuscript: NoDM, Increased Insulin Resistance, Midzone Glycemia, Prediabetes, Diabetes

Use in manuscript: Phenotype assignment only; not used as an adaptive diagnostic, predictive, or machine-learning system

The archived calculator package should include the static HTML/JavaScript source file, phenotype-classification logic, reference thresholds used by the algorithm, and the output-category mapping used for the present analysis.

### S1.5 Reproducibility statement

The exact phenotype classification rules and the analysis-locked calculator version were archived before manuscript submission and provided as Supplementary Methods. This was done to ensure that the OGTT-derived phenotype assignment can be independently reviewed, reproduced, and distinguished from any later modifications to the public-facing calculator.

